# Clinical Validation of Segmentation-Based Detection of Glioma Progression

**DOI:** 10.1101/2022.08.17.22278562

**Authors:** Pablo F. Damasceno, Tyler Gleason, James Hawkins, Tracy Luks, Sharmila Majumdar, Janine M. Lupo, Jason C. Crane, Javier E. Villanueva-Meyer

## Abstract

**Purpose:** To evaluate whether an AI-based method could be used routinely as part of patient care to assist in detecting non-enhancing glioma progression.

**Materials and Methods:** A 3D U-Net trained (n=481) and validated (n=121) to segment post-surgical lower grade gliomas was used to measure tumor volumes over time and assess progression in a clinical test set. Eight prospective and eight retrospective patients (total 72 exams) who were suspected of progression during their routine outpatient imaging were clinically assessed. Gold standards for progression were derived from clinical reports a posteriori using visual read, and radiologists were blinded to the AI decision at time of reporting.

**Results:** Progression assessments were presented to radiologists via an easy-to-use, interactive, and interpretable environment in under 10 minutes. Combining prospective and retrospective cases, a final sensitivity of 0.72 and specificity of 0.75 was achieved at progression detection.

**Conclusions:** Automated detection of glioma progression would provide valuable decision support for routine use.

## Introduction

Radiological assessment of treatment response of glioma is primarily based on change of tumor size on anatomical MRI sequences. Subtle increases in size can be difficult to detect on serial MRI exams, potentially leading to delays in treatment^1–3^. Although assessment of tumor progression is known to have smaller variance when performed volumetrically^4^, the intensive human labor involved in segmentation limits its routine use in clinical practice^5^.

Deep learning methods are particularly suited for medical image evaluation tasks and have been shown to perform well in multiple neuroimaging contexts^6^. However, most reports of AI-informed models focus on performance accuracy, and often do not address the many challenges associated with clinical deployment, such as data quality and variability, usability in clinical workflows, the handling of errors, and transparent presentation of clinically useful and interpretable results ^7^.

To bridge the gap from research to clinically deployable AI, a lesion segmentation model to assist with the automated detection of tumor progression in lower grade gliomas was developed and deployed for clinical validation. The model was trained on T2-weighted FLAIR images from post-surgical cases, which are of particular importance because resection can lead to brain shifts and image artifacts that make it difficult for automated segmentation algorithms to correctly delineate the tumor^8^.

Preliminary results of the clinical validation of our model on accurately detecting glioma progression are presented here to highlight the challenges that must be overcome prior to adoption of a model for routine use in clinical care. The method is based on real-time, neural-network-based lesion segmentation and longitudinal changes in associated volumetrics to predict progression. By integrating automatic image pre-processing, fast and accurate segmentation, and a deployment pipeline for model visualization and evaluation, 3D changes in glioma volume can be tracked over time and presented in an interpretable way to the radiologist.

### Project goal and software design

The primary goal of this project was to evaluate whether an AI-based method could be used routinely as part of patient care to assist in detecting non-enhancing glioma progression. The software package was intended to be: i) easy to use, showing minimal impact on radiologists’ workflow; ii) interactive, offering the ability for real-time choice of baseline exam and threshold for progression; and iii) interpretable, showing 3D image segmentations side-by-side with tumor volumes to facilitate interpretation of the model results. For that reason, both the number of times the AI-derived progression agreed with radiologist reports as well as the radiologists’ sentiment on software usability were used as success metrics.

## Materials and Methods

### Data collection

The study received institutional review board approval with consent waiver. 605 T2-weighted FLAIR images from a single institution (GE 750 3T scanner, 3D sagittal acquisitions) were split into training/test sets (80%, 20%). Manually or semi-automatically segmented T2-hyperintense lesions were defined on FLAIR images by experienced investigators using open-source software (3D Slicer^9^). Automatic segmentation of lesion volumes was performed using an encoder-decoder architecture^10^ previously found to perform well on pre-surgical brain tumor cases^11^. Pre-processing and post-processing steps including data augmentation were performed as described by Myronenko^10^, modified for single channel (FLAIR), rather than four channel (FLAIR, T1 pre-contrast, T1 post-contrast, T2) input.

### Automatic segmentation and clinical integration

Clara Deploy^12^ was used as the inference service for the model and to handle the communication between PACS and an XNAT^13^ server where results were stored and displayed. Selected exams being read in PACS were sent in real time to a virtual machine running Clara Deploy which identified the desired FLAIR series, performed the lesion segmentation, and stored the results in XNAT. Radiologists could then log into a dedicated XNAT webpage where cases were displayed in tabular form together with respective tumor volumes and progression assessment. In-browser visualization of images and AI-derived segmentations was supported by OHIF^14^ (Figure 1).

**Figure 1.**
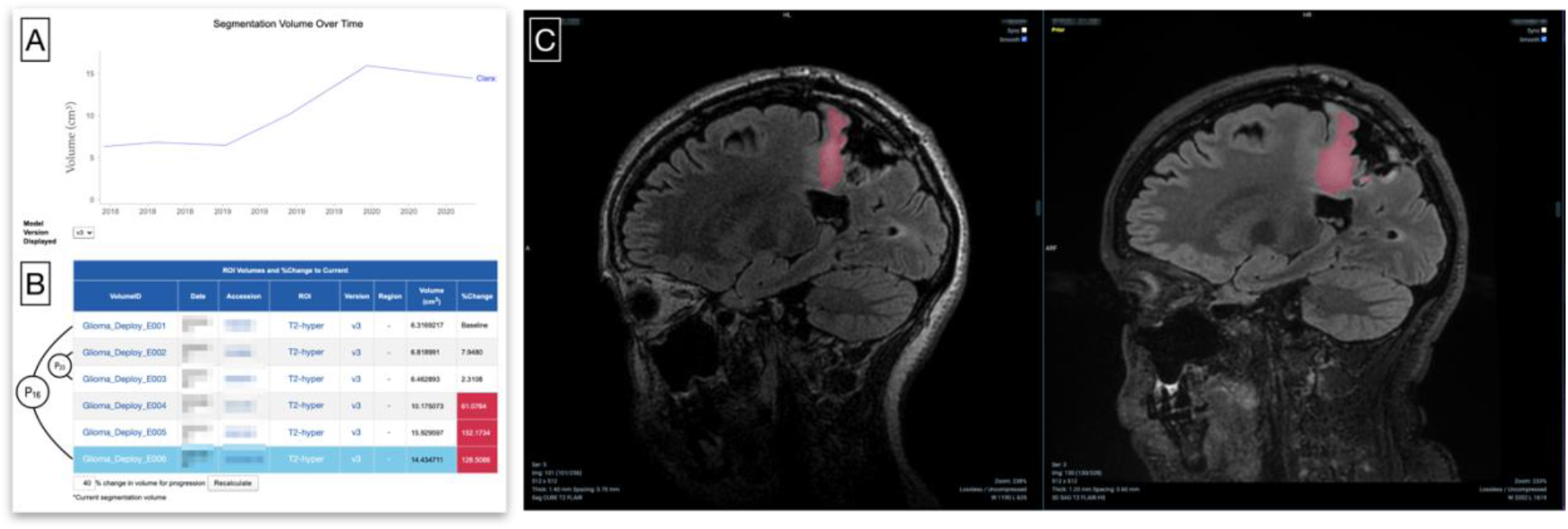
Deployed tumor progression detection pipeline in action. Visual representation of glioma volume over time (A) followed by a table of individual exams (B) containing tumor volumes, and percentage change with respect to baseline (highlighted in blue). Arrow connections exemplify two *baseline-to-follow-up* pairs. Exams whose tumor volume exceeds a chosen threshold (defaulted to 40%) are marked in red to indicate progression. Interactive options include: i) selection of AI model version; ii) selection of baseline exam; iii) manipulation of threshold percentage value; and iv) ability to choose which exam to be visualized. (C) in-browser visualization of two exams – baseline (left) and latest (right) – as well as AI-generated glioma segmentation (pink). The volume increase is clear and facilitates the visual confirmation of progression or identification of model mistakes.

### Model interpretability

Instead of directly modeling progression, image segmentation and volume change heuristics for assessing progression were employed to provide clinicians with a transparent decision process where thresholds, baseline exams, and segmentations are selected and editable at will Figure 1). This was intended to enhance confidence in results by exposing the visually interpretable segmentations underlying the progression predictions.

### Progression detection heuristic

Tumors were marked as ‘progressed’ for volume changes larger than 40% between baseline and the MRI of interest^1^. Because the baseline exam was variable based on treatment, the system allowed the clinician to choose the baseline scan interactively. This allowed, for instance, selection of baseline tumors with volume greater than 1cm^3^, as suggested by van den Bent et al^1^. Once a baseline exam was chosen, all other exams were marked as blue (stable) or red (progressed) with respect to that baseline (Figure 1).

### Prospective evaluation

Eight prospective cases were obtained during a two-week period from patients who were suspected of progression during routine outpatient imaging. The current MRI and two preceding MRIs (24 total exams) were manually pushed from PACS to the AI algorithm for segmentation. Gold standards for progression were derived from clinical reports *a posteriori* using visual read, and radiologists were blinded to the AI decision at the time of their reporting.

### Retrospective evaluation

To evaluate model true and false positive rates, cases with slow progression were identified via text search of radiology reports. In total, 8 patients with 6 visits each for a total of 48 exams were analyzed.

## Results

### Segmentation training

The model achieved a mean DICE score of 0.87 ± 0.20 for AI-segmented FLAIR lesions on 124 images composing the test set. The 90th percentile DICE score was also 0.87, revealing that most images had an excellent agreement with ground truth.

### Prospective evaluation

Radiology reports revealed that all 24 baseline-to-follow-up **pairs were marked as stable, of which 20 were correctly identified as such by the** algorithm, resulting in a specificity = 0.83 (see Fig. 2A-B).

**Figure 2.**
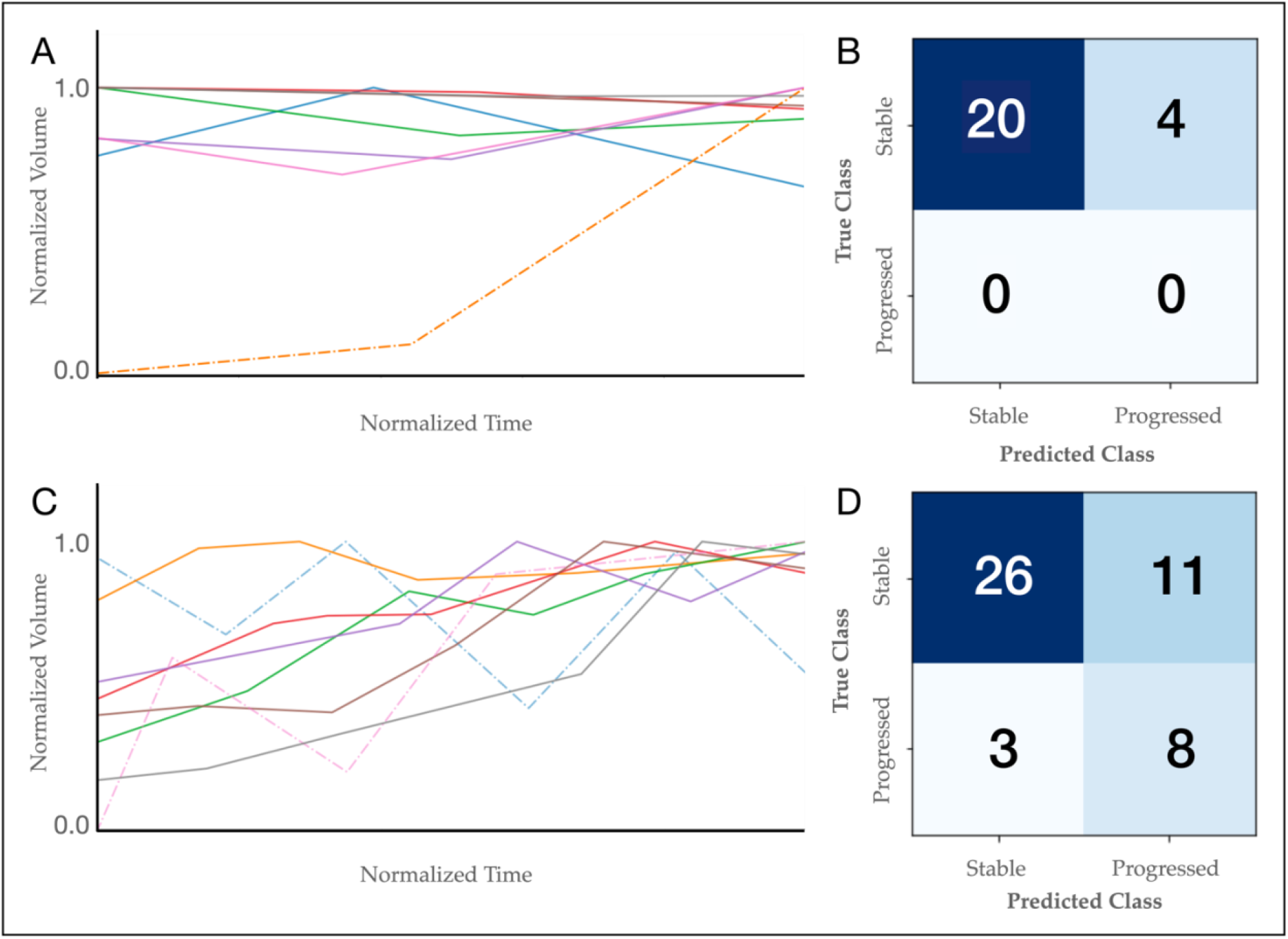
Glioma segmentation volumes and progression detection. Glioma volumes as a function of time for prospective (A) and retrospective (C) cases. Dashed lines show cases where the model made significant mistakes due to acquisition protocol issues. Volumes were normalized by the maximum volume observed to facilitate collected visualization.

### Retrospective evaluation

For the 48 exams analyzed (8 patients, 6 serial exams per patient), 26 out of the total 37 stable cases were correctly identified as such by the model upon correct choice of baseline, for a specificity of 0.70. For the progression cases, 8 out of 11 cases were correctly identified, for a sensitivity of 0.72 (Fig. 2C-D).

### Model performance

The automated volumetrics measurement showed variance between 2-18% which, for all these cases, is below the 9 mL variability observed in manual diameter calculation^15^. For all but one patient, AI progression assessment discrepancies with gold standard were due to imaging acquisitions differing significantly from those used during model training (GE 3D Cube). For the remaining case, a small increase in tumor volume from 1.7 mL to 2.7 mL was likely below the threshold for human reader detection^4,15^ and only later marked as progressive.

### System Usability

All prospective results were available for the radiologist within 10 minutes of DICOM transmission (90th percentile: 4 minutes and 40 seconds). 72 out of 73 cases were processed correctly (success rate: 99%). Two radiologists reported that the results and presentation were interpretable and would provide valuable decision support for routine use if data selection could be automated.

## Discussion and Conclusions

This work presents the first steps toward clinical deployment of an AI-based solution for real-time detection of glioma progression. Combining prospective and retrospective cases, a final sensitivity of 0.72 and specificity of 0.75 was achieved. 90% of the successfully processed cases were available in under 5 minutes and only one case in 73 failed to be processed. Results were presented via an easy-to-use, interactive, and explainable environment. Radiologists could manually choose baseline cases, adjust threshold for progression, visually inspect segmentations associated with the exam, and graphically check how volume varied over time.

Discrepancies between model and ground truth were mostly due to imaging acquisitions differing from those used during model training (Fig. 3). This should be mitigated through additional training iterations on a more representative clinical cohort, with varied acquisition parameters. Additionally, if inadequate data does arrive at the AI model, the output should explicitly note that the model confidence is low as opposed to simply giving the wrong answer.

**Figure 3.**
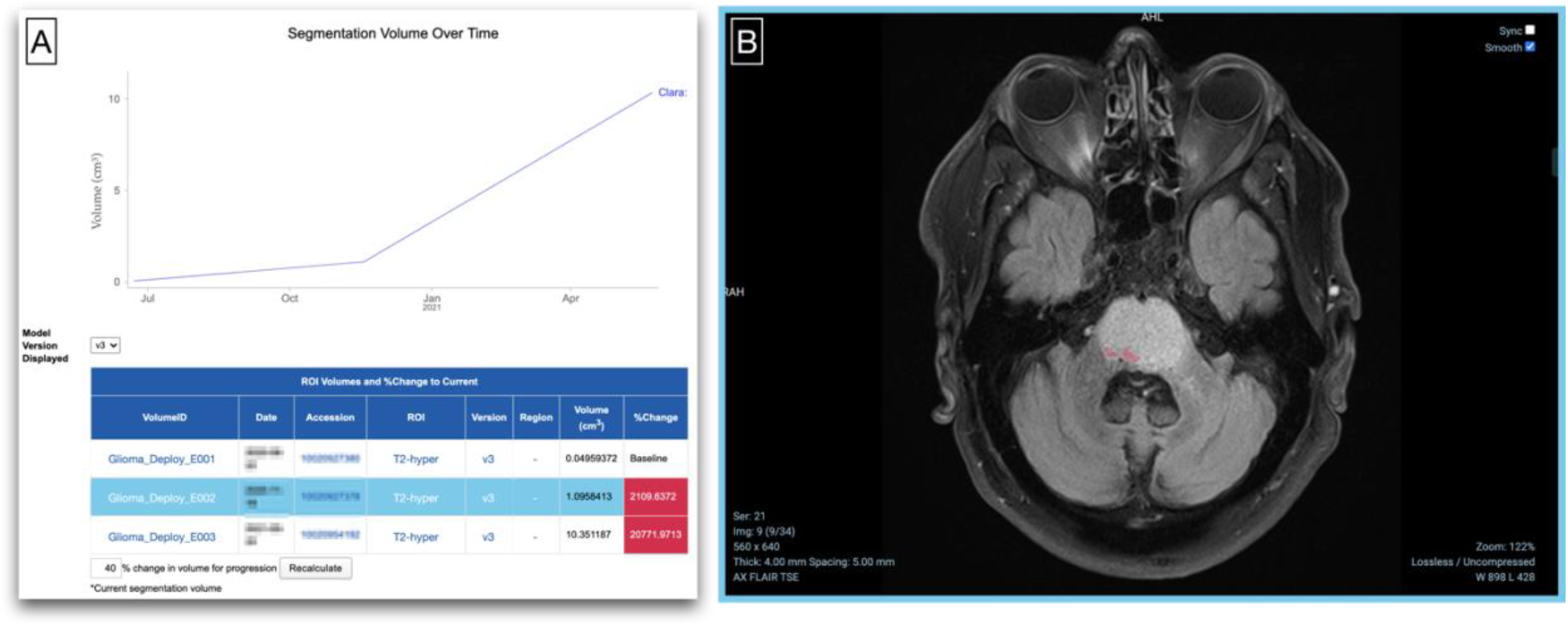
An example of an AI tumor progression misclassification. (A) Glioma volume over time for a patient whose prior visits came from another institution. Because the prior data was a 2D acquisition – and therefore different from what the model was trained on – the AI model was not able to segment the glioma clearly present in the image (B).

A potential solution for both issues would involve a “human in the loop” scenario, where images would be first sent to a dedicated QC team responsible for checking (input / output) data quality prior to notifying the doctor about the results. This would yield a database of common ‘real-life’ failures that future models could be trained on. Additionally, QC team triage would mitigate the risk of unintended clinical consequences^7^ and increase trust from clinicians and patients.

The aforementioned performance nevertheless highlights the benefit of the automated method to reduce variability in FLAIR lesion measurement and hence increase accuracy in disease progression detection. To maximize applicability at all treatment timepoints, the ability to change the baseline provides a needed function particularly in the context of changing therapies or clinical trials.

## Data Availability

Data that support the findings of this study are available from the corresponding author upon reasonable request.

